# Risk of myocarditis and pericarditis following BNT162b2 and ChAdOx1 COVID-19 vaccinations

**DOI:** 10.1101/2022.03.06.21267462

**Authors:** Samantha Ip, Fatemeh Torabi, Spiros Denaxas, Ashley Akbari, Hoda Abbasizanjani, Rochelle Knight, Jennifer Cooper, Rachel Denholm, Spencer Keene, Thomas Bolton, Sam Hollings, Efosa Omigie, Teri-Louise North, Arun Karthikeyan Suseeladevi, Emanuele Di Angelantonio, Kamlesh Khunti, Jonathan A C Sterne, Cathie Sudlow, William Whiteley, Angela Wood, Venexia Walker, the British Heart Foundation Data Science Centre (Health Data Research UK) CVD-COVID-UK/COVID-IMPACT Consortium and the Longitudinal Health and Wellbeing and Data and Connectivity UK COVID-19 National Core Studies

## Abstract

We describe our analyses of data from over 49.7 million people in England, representing near-complete coverage of the relevant population, to assess the risk of myocarditis and pericarditis following BNT162b2 and ChAdOx1 COVID-19 vaccination. A self-controlled case series (SCCS) design has previously reported increased risk of myocarditis after first ChAdOx1, BNT162b2, and mRNA-1273 dose and after second doses of mRNA COVID-19 vaccines in England. Here, we use a cohort design to estimate hazard ratios for hospitalised or fatal myocarditis/pericarditis after first and second doses of BNT162b2 and ChAdOx1 vaccinations. SCCS and cohort designs are subject to different assumptions and biases and therefore provide the opportunity for triangulation of evidence. In contrast to the findings from the SCCS approach previously reported for England, we found evidence for lower incidence of hospitalised or fatal myocarditis/pericarditis after first ChAdOx1 and BNT162b2 vaccination, as well as little evidence to suggest higher incidence of these events after second dose of either vaccination.

## Main text

On May 17th 2021, the US Centers for Disease Control and Prevention (CDC) reported mild to moderate myocarditis/pericarditis cases following a second dose mRNA COVID-19 vaccinations. Most patients were men <50 years old, presenting with chest pain, fever and raised cardiac-specific troponin several days after vaccination, followed by full recovery.^1^ A higher risk of myocarditis, particularly in younger men, was reported from Israel following second BNT162b2 vaccination.^2^ In England, a self-controlled case series (SCCS) design reported increased risks of myocarditis after first ChAdOx1, BNT162b2, and mRNA-1273 dose and after second doses of mRNA COVID-19 vaccines.^3^ Here, we use a cohort design to estimate adjusted hazard ratios (aHRs) for hospitalised or fatal myocarditis/pericarditis after first and second BNT162b2 and ChAdOx1 vaccinations. SCCS and cohort designs are subject to different assumptions and biases, so these results provide an opportunity for triangulation of evidence.

We analysed linked national electronic health records from multiple sources in England, including primary and secondary care, death registries, COVID-19 testing and vaccination data.^4,5^ Our analyses included up to 49,786,346 people aged 12+ years, of known sex, and alive at the start of the vaccination programme (December 8th 2020) with at least one primary care record. This corresponds to near-complete coverage of the relevant population. The outcomes ‘myocarditis’ and ‘pericarditis’ were defined using ICD10 codes (myocarditis: I40*,I41*,I51.4; pericarditis: I30*) recorded in any position for a hospital admission or cause of death. Outcome date was taken as the date of first relevant code and the combined outcome ‘myocarditis/pericarditis’ used the earliest date of either outcome. Follow-up began on December 8th 2020 for first dose analyses and on date of first vaccination for second dose analyses. Follow-up ended with the earliest of: outcome event, death, or study end date (May 17^th^ 2021). Additionally, follow-up for first dose analyses ended at second vaccination with the same product or receipt of a non-BNT162b2 (for the BNT162b2 analysis) or non-ChAdOx1-S (for the ChAdOx1-S analysis) vaccination.

We estimated aHRs for hospitalised or fatal myocarditis/pericarditis 0-13 and 14+ days after first and second BNT162b2 and ChAdOx1-S vaccination using Cox models to account for calendar time. We adjusted for age, sex, region, socioeconomic status, ethnicity, prior COVID-19, and prior myocarditis/pericarditis. We examined interactions with sex and age group. For computational efficiency, we included all people with the outcome and a 10% random sample without the outcome. Inverse probability weights were used to account for sampling and confidence intervals derived using robust standard errors. To avoid ascertainment bias, follow-up ended on May 17th 2021, because clinicians aware of CDC reports may have lowered diagnostic thresholds in vaccinated compared with unvaccinated patients after that date. During this period, individuals aged 38+ years, along with the clinically vulnerable and frontline health and social care workers of any age, were eligible for vaccination. We replicated first dose analyses in Wales (N = 2,615,853) but had insufficient events to replicate other analyses. We performed two sensitivity analyses: (1) subgroup analysis in people with and without prior COVID-19 and (2) analysis of ‘myocarditis’ and ‘pericarditis’ separately collapsed into a single post-vaccination period. Our pre-specified protocol, code and additional analyses are publicly available (https://github.com/BHFDSC/CCU002_03).

We identified 13,216,259 and 8,614,934 people who received first and second doses of BNT162b2; and 19,336,909 and 15,925,122 who received first and second doses of ChAdOx1. There were 1,459 events pre- and 607 events post-vaccination. We found lower incidence of myocarditis/pericarditis after first vaccination (0-13 days aHRs: BNT162b2 0.58, 95% CI 0.39-0.84; ChAdOx1-S 0.67, 95% CI 0.50-0.89, 14+ days aHRs: BNT162b2 0.80, 95% CI 0.65-0.97; ChAdOx1-S 0.75, 95% CI 0.63-0.90) compared with before or without vaccination. After second BNT162b2 vaccination the aHRs were: 0-13 days 0.77 (95% CI 0.50-1.18) and 14+ days 1.11 (95% CI 0.79-1.55) compared with the period between first and second doses. Corresponding aHRs following second ChAdOx1-S vaccination were 1.33 (95% CI 0.84-2.10) and 1.38 (95% CI 0.85-2.24). There was inconsistent evidence for interactions by sex and age group for second dose analyses (all heterogeneity p-values>=0.1), with low event counts preventing clear inferences. However, the aHRs for first dose analyses decreased with increasing age group. Subgroup analysis of people with and without prior COVID-19 produced similar aHRs. Splitting the combined outcome suggested that pericarditis may be driving elevated aHRs (e.g., second ChAdOx1-S vaccination: pericarditis 2.04, 95% CI 1.12-3.70; myocarditis 1.24, 95% CI 0.75-2.04).

In contrast to the SCCS approach findings, we found evidence for lower incidence of hospitalised or fatal myocarditis/pericarditis after first ChAdOx1 and BNT162b2 vaccination. (3) Furthermore, we found little evidence to suggest higher incidence of these events after second dose of either vaccination. Myocarditis/pericarditis is rare after second BNT162b2 vaccination. Benefits of COVID-19 vaccination therefore substantially outweigh the risk of myocarditis/pericarditis.

## Data Availability

The data used in this study are available in NHS Digital's TRE for England, but as restrictions apply they are not publicly available (https://digital.nhs.uk/coronavirus/coronavirus-data-services-updates/trusted- research-environment-service-for-england). The CVD-COVID-UK/COVID-IMPACT programme led by the BHF Data Science Centre (https://www.hdruk.ac.uk/helping-with-health-data/bhf-data-science-centre/) received approval to access data in NHS Digital's TRE for England from the Independent Group Advising on the Release of Data (IGARD) (https://digital.nhs.uk/about-nhs-digital/corporate-information-and-documents/independent-group-advising-on-the-release-of-data) via an application made in the Data Access Request Service (DARS) Online system (ref. DARS-NIC-381078-Y9C5K) (https://digital.nhs.uk/services/data-access-request-service-dars/dars-products-and-services). The CVD-COVID-UK/COVID-IMPACT Approvals & Oversight Board (https://www.hdruk.ac.uk/projects/cvd-covid-uk-project/) subsequently granted approval to this project to access the data within the TRE for England and the Secure Anonymised Information Linkage (SAIL) Databank. The de-identified data used in this study was made available to accredited researchers only.
The data used in this study are available in the SAIL Databank at Swansea University, Swansea, UK, but as restrictions apply they are not publicly available. All proposals to use SAIL data are subject to review by an independent Information Governance Review Panel (IGRP). Before any data can be accessed, approval must be given by the IGRP. The IGRP gives careful consideration to each project to ensure proper and appropriate use of SAIL data. When access has been granted, it is gained through a privacy protecting safe haven and remote access system referred to as the SAIL Gateway. SAIL has established an application process to be followed by anyone who would like to access data via SAIL at https://www.saildatabank.com/application-process.

---

**Figure:**
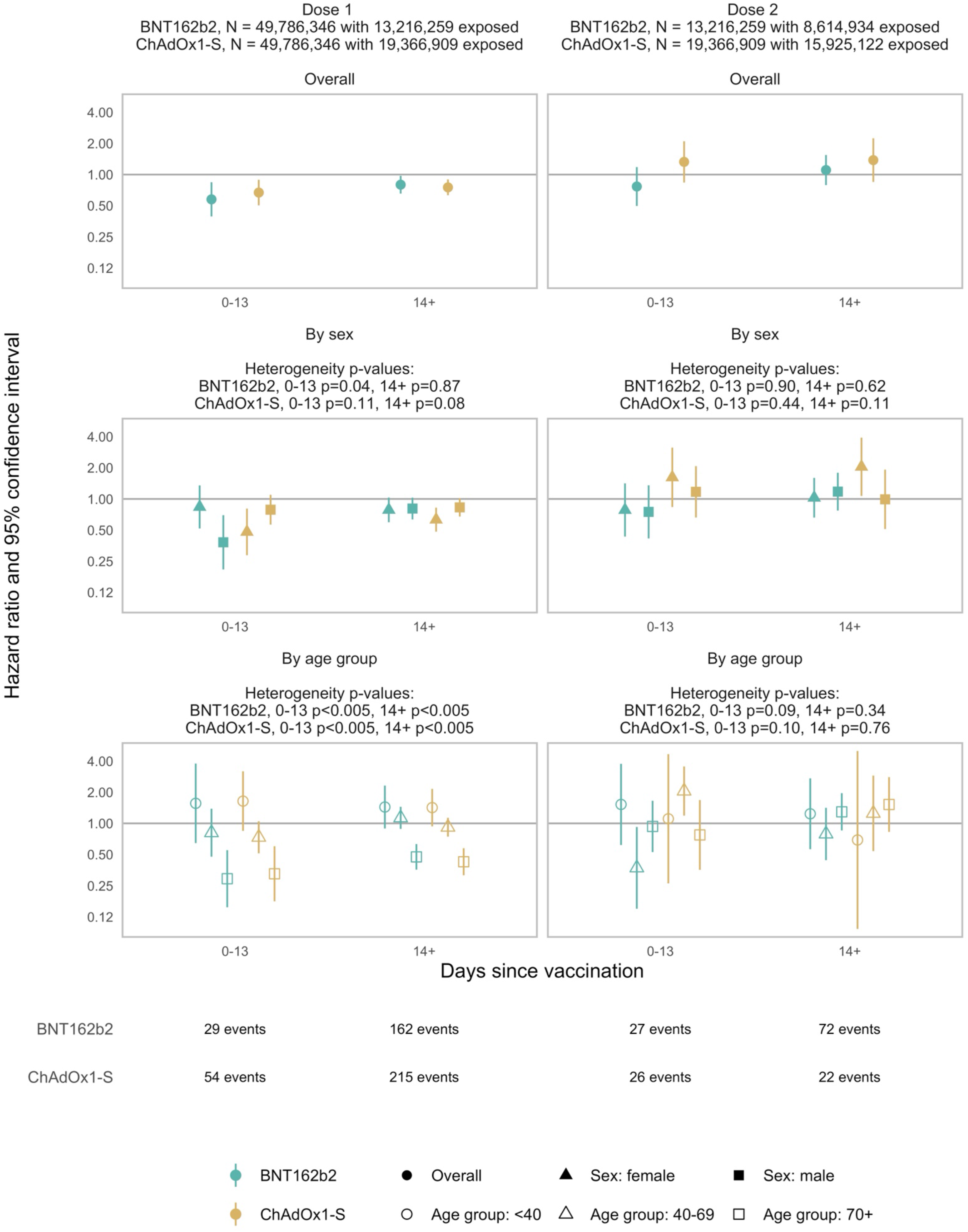
Hazard ratio and 95% confidence interval for myocarditis/pericarditis following each dose of vaccine, by vaccine type.

