## Supplement for "Risk of myocarditis and pericarditis following BNT162b2 and ChAdOx1 COVID-19 vaccinations"

### Sample size

#### Dose 1

| analysis_product | sample | N |
|---|---|---|
| BNT162b2 | Exposed | 13216259 |
| BNT162b2 | Total | 49786346 |
| ChAdOx1-S | Exposed | 19366909 |
| ChAdOx1-S | Total | 49786346 |

#### Dose 2

| analysis_product | sample | N |
|---|---|---|
| BNT162b2 | Exposed | 8614934 |
| BNT162b2 | Total | 13216259 |
| ChAdOx1-S | Exposed | 15925122 |
| ChAdOx1-S | Total | 19366909 |

#### Patient characteristics

| Category | Sub-category | BNT162b2 | | | ChAdOx1 | | |
|---|---|---|---|---|---|---|---|
| Age Group | | <40 | 40-69 | 70+ | <40 | 40-69 | 70+ |
| N | | 5285006 | 4412060 | 3519193 | 2683141 | 13368016 | 3315752 |
| Sex | Male | 2487435 | 1900760 | 1594618 | 1135001 | 6833559 | 1478295 |
| Sex | Female | 2797571 | 2511300 | 1924575 | 1548140 | 6534457 | 1837457 |
| Ethnicity | Asian or Asian British | 549040 | 386154 | 108015 | 308781 | 908142 | 98539 |
| Ethnicity | Black or Black British | 111271 | 141237 | 33590 | 76271 | 367260 | 39489 |
| Ethnicity | Mixed | 94880 | 50062 | 11442 | 50015 | 134397 | 12033 |
| Ethnicity | Other Ethnic Groups | 171014 | 86458 | 26778 | 69486 | 268767 | 26373 |
| Ethnicity | White | 4190348 | 3670391 | 3294163 | 2129790 | 11350894 | 3089673 |
| Ethnicity | Unknown Or Missing | 168453 | 77758 | 45205 | 48798 | 338556 | 49645 |
| Deprivation | Quintile 1 (most deprived) | 881344 | 790045 | 443962 | 572438 | 2207082 | 461548 |
| Deprivation | Quintile 2 | 981813 | 835141 | 581166 | 534701 | 2419578 | 556935 |
| Deprivation | Quintile 3 | 961616 | 881695 | 731166 | 477660 | 2615004 | 678834 |
| Deprivation | Quintile 4 | 927203 | 887068 | 817091 | 436929 | 2739606 | 736807 |
| Deprivation | Quintile 5 (least deprived) | 871970 | 867543 | 863689 | 390149 | 2857595 | 759426 |
| History | Coronavirus Infection | 81731 | 160404 | 59623 | 92381 | 285426 | 77470 |
| History | Myocarditis/pericarditis | 3434 | 5551 | 4046 | 2975 | 13069 | 3843 |
| Region | North West | 617093 | 608345 | 468799 | 345748 | 1646966 | 396372 |
| Region | Missing | 950906 | 438841 | 331880 | 412244 | 1431009 | 385804 |
| Region | London | 782420 | 624630 | 361282 | 389359 | 1682984 | 248603 |
| Region | South East | 685750 | 623726 | 560464 | 332962 | 2092038 | 537598 |
| Region | East Of England | 428282 | 376427 | 355180 | 221443 | 1284366 | 324343 |
| Region | South West | 372983 | 377451 | 333654 | 165922 | 1104950 | 331338 |
| Region | East Midlands | 357990 | 314067 | 282392 | 195491 | 989434 | 249357 |
| Region | Yorkshire And The Humber | 461281 | 419032 | 326902 | 257455 | 1299522 | 351528 |
| Region | West Midlands | 452944 | 446357 | 369992 | 266374 | 1315953 | 346754 |
| Region | North East | 175357 | 183184 | 128648 | 96143 | 520794 | 144055 |

### Patient age distribution

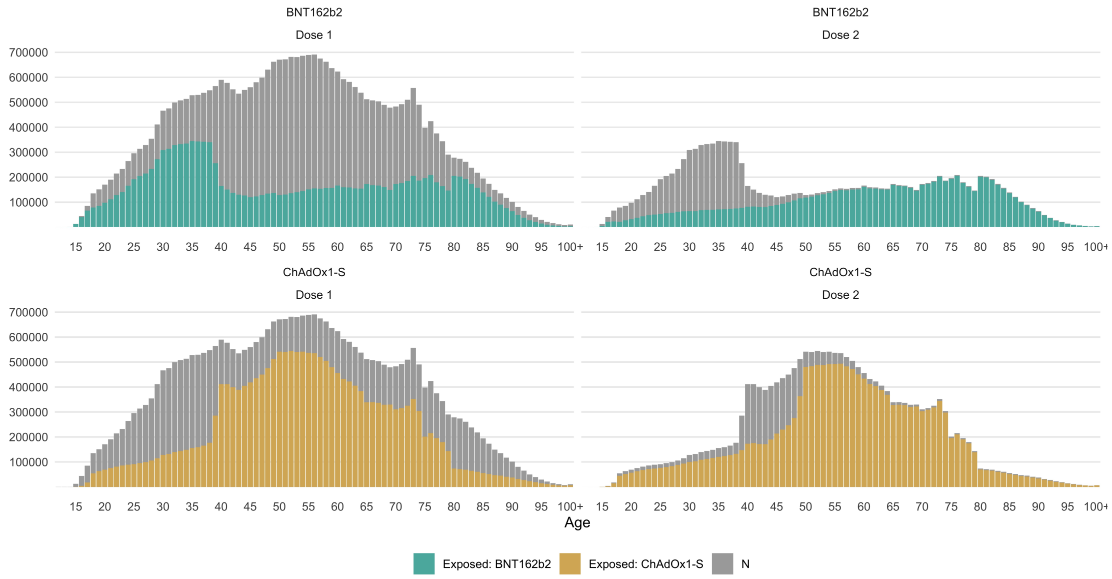

### Estimates, overall

#### Dose 1

| exposure | days_post_vaccination | estimate | conf.low | conf.high | p.value |
|---|---|---|---|---|---|
| BNT162b2 | 0-13 | 0.58 | 0.39 | 0.84 | 0.00 |
| BNT162b2 | 14+ | 0.80 | 0.65 | 0.97 | 0.03 |
| ChAdOx1-S | 0-13 | 0.67 | 0.50 | 0.89 | 0.01 |
| ChAdOx1-S | 14+ | 0.75 | 0.63 | 0.90 | 0.00 |

#### Dose 2

| exposure | days_post_vaccination | estimate | conf.low | conf.high | p.value |
|---|---|---|---|---|---|
| BNT162b2 | 0-13 | 0.77 | 0.50 | 1.18 | 0.23 |
| BNT162b2 | 14+ | 1.11 | 0.79 | 1.55 | 0.56 |
| ChAdOx1-S | 0-13 | 1.33 | 0.84 | 2.10 | 0.23 |
| ChAdOx1-S | 14+ | 1.38 | 0.85 | 2.24 | 0.20 |

### Estimates, by sex

#### Dose 1

| exposure | days_post_vaccination | sex | estimate | conf.low | conf.high | p.value |
|---|---|---|---|---|---|---|
| BNT162b2 | 0-13 | Male | 0.38 | 0.21 | 0.70 | 0.01 |
| BNT162b2 | 0-13 | Female | 0.84 | 0.52 | 1.35 | 0.92 |
| BNT162b2 | 14+ | Male | 0.81 | 0.63 | 1.03 | 0.30 |
| BNT162b2 | 14+ | Female | 0.78 | 0.60 | 1.03 | 0.29 |
| ChAdOx1-S | 0-13 | Male | 0.79 | 0.56 | 1.10 | 0.49 |
| ChAdOx1-S | 0-13 | Female | 0.48 | 0.29 | 0.81 | 0.02 |
| ChAdOx1-S | 14+ | Male | 0.83 | 0.68 | 1.02 | 0.25 |
| ChAdOx1-S | 14+ | Female | 0.63 | 0.48 | 0.82 | 0.00 |

#### Dose 2

| exposure | days_post_vaccination | sex | estimate | conf.low | conf.high | p.value |
|---|---|---|---|---|---|---|
| BNT162b2 | 0-13 | Male | 0.75 | 0.42 | 1.35 | 0.80 |
| BNT162b2 | 0-13 | Female | 0.78 | 0.43 | 1.41 | 0.88 |
| BNT162b2 | 14+ | Male | 1.18 | 0.77 | 1.79 | 0.90 |
| BNT162b2 | 14+ | Female | 1.03 | 0.66 | 1.60 | 1.00 |
| ChAdOx1-S | 0-13 | Male | 1.17 | 0.66 | 2.08 | 0.97 |
| ChAdOx1-S | 0-13 | Female | 1.62 | 0.83 | 3.13 | 0.48 |
| ChAdOx1-S | 14+ | Male | 0.99 | 0.51 | 1.92 | 1.00 |
| ChAdOx1-S | 14+ | Female | 2.05 | 1.07 | 3.91 | 0.12 |

#### Estimates, by age group

##### Dose 1

| exposure | days_post_vaccination | age_group | estimate | conf.low | conf.high | p.value |
|---|---|---|---|---|---|---|
| BNT162b2 | 0-13 | <40 | 1.56 | 0.64 | 3.78 | 0.90 |
| BNT162b2 | 0-13 | 40-69 | 0.81 | 0.48 | 1.39 | 0.97 |
| BNT162b2 | 0-13 | 70+ | 0.29 | 0.15 | 0.55 | 0.00 |
| BNT162b2 | 14+ | <40 | 1.44 | 0.89 | 2.32 | 0.58 |
| BNT162b2 | 14+ | 40-69 | 1.13 | 0.88 | 1.45 | 0.91 |
| BNT162b2 | 14+ | 70+ | 0.47 | 0.36 | 0.63 | 0.00 |
| ChAdOx1-S | 0-13 | <40 | 1.64 | 0.85 | 3.18 | 0.60 |
| ChAdOx1-S | 0-13 | 40-69 | 0.73 | 0.51 | 1.05 | 0.42 |
| ChAdOx1-S | 0-13 | 70+ | 0.33 | 0.18 | 0.60 | 0.00 |
| ChAdOx1-S | 14+ | <40 | 1.42 | 0.94 | 2.15 | 0.47 |
| ChAdOx1-S | 14+ | 40-69 | 0.92 | 0.74 | 1.13 | 0.96 |
| ChAdOx1-S | 14+ | 70+ | 0.43 | 0.32 | 0.57 | 0.00 |

##### Dose 2

| exposure | days_post_vaccination | age_group | estimate | conf.low | conf.high | p.value |
|---|---|---|---|---|---|---|
| BNT162b2 | 0-13 | <40 | 1.52 | 0.62 | 3.76 | 0.93 |
| BNT162b2 | 0-13 | 40-69 | 0.37 | 0.15 | 0.92 | 0.18 |
| BNT162b2 | 0-13 | 70+ | 0.93 | 0.53 | 1.65 | 1.00 |
| BNT162b2 | 14+ | <40 | 1.24 | 0.56 | 2.72 | 1.00 |
| BNT162b2 | 14+ | 40-69 | 0.79 | 0.44 | 1.41 | 0.96 |
| BNT162b2 | 14+ | 70+ | 1.29 | 0.85 | 1.96 | 0.77 |
| ChAdOx1-S | 0-13 | <40 | 1.11 | 0.26 | 4.66 | 1.00 |
| ChAdOx1-S | 0-13 | 40-69 | 2.05 | 1.19 | 3.54 | 0.06 |
| ChAdOx1-S | 0-13 | 70+ | 0.77 | 0.36 | 1.68 | 0.99 |
| ChAdOx1-S | 14+ | <40 | 0.69 | 0.10 | 5.00 | 1.00 |
| ChAdOx1-S | 14+ | 40-69 | 1.25 | 0.54 | 2.89 | 1.00 |
| ChAdOx1-S | 14+ | 70+ | 1.52 | 0.83 | 2.80 | 0.69 |

### Heterogeneity p-values

#### Dose 1

##### Sex

| exposure | days_post_vaccination | p.value |
|---|---|---|
| BNT162b2 | 0-13 | 0.06 |
| BNT162b2 | 0-13 | <0.005 |
| BNT162b2 | 0-13 | 0.04 |
| BNT162b2 | 14+ | 0.80 |
| BNT162b2 | 14+ | 0.62 |
| BNT162b2 | 14+ | 0.87 |
| ChAdOx1-S | 0-13 | 0.15 |
| ChAdOx1-S | 0-13 | 0.46 |
| ChAdOx1-S | 0-13 | 0.11 |
| ChAdOx1-S | 14+ | 0.06 |
| ChAdOx1-S | 14+ | 0.65 |
| ChAdOx1-S | 14+ | 0.08 |

##### Age group

| exposure | days_post_vaccination | p.value |
|---|---|---|
| BNT162b2 | 0-13 | <0.005 |
| BNT162b2 | 0-13 | <0.005 |
| BNT162b2 | 0-13 | <0.005 |
| BNT162b2 | 14+ | <0.005 |
| BNT162b2 | 14+ | <0.005 |
| BNT162b2 | 14+ | <0.005 |
| ChAdOx1-S | 0-13 | <0.005 |
| ChAdOx1-S | 0-13 | <0.005 |
| ChAdOx1-S | 0-13 | <0.005 |
| ChAdOx1-S | 14+ | <0.005 |
| ChAdOx1-S | 14+ | 0.45 |
| ChAdOx1-S | 14+ | <0.005 |

#### Dose 2

##### Sex

| exposure | days_post_vaccination | p.value |
|---|---|---|
| BNT162b2 | 0-13 | 0.89 |
| BNT162b2 | 0-13 | 0.89 |
| BNT162b2 | 0-13 | 0.90 |
| BNT162b2 | 14+ | 0.94 |
| BNT162b2 | 14+ | 0.94 |
| BNT162b2 | 14+ | 0.62 |
| ChAdOx1-S | 0-13 | 0.55 |
| ChAdOx1-S | 0-13 | 0.55 |
| ChAdOx1-S | 0-13 | 0.44 |
| ChAdOx1-S | 14+ | 0.23 |
| ChAdOx1-S | 14+ | 0.23 |
| ChAdOx1-S | 14+ | 0.11 |

##### Age group

| exposure | days_post_vaccination | p.value |
|---|---|---|
| BNT162b2 | 0-13 | 0.07 |
| BNT162b2 | 0-13 | 0.07 |
| BNT162b2 | 0-13 | 0.09 |
| BNT162b2 | 14+ | 0.27 |
| BNT162b2 | 14+ | 0.27 |
| BNT162b2 | 14+ | 0.34 |
| ChAdOx1-S | 0-13 | 0.11 |
| ChAdOx1-S | 0-13 | 0.11 |
| ChAdOx1-S | 0-13 | 0.10 |
| ChAdOx1-S | 14+ | 0.51 |
| ChAdOx1-S | 14+ | 0.51 |
| ChAdOx1-S | 14+ | 0.76 |

#### Estimates, by prior history

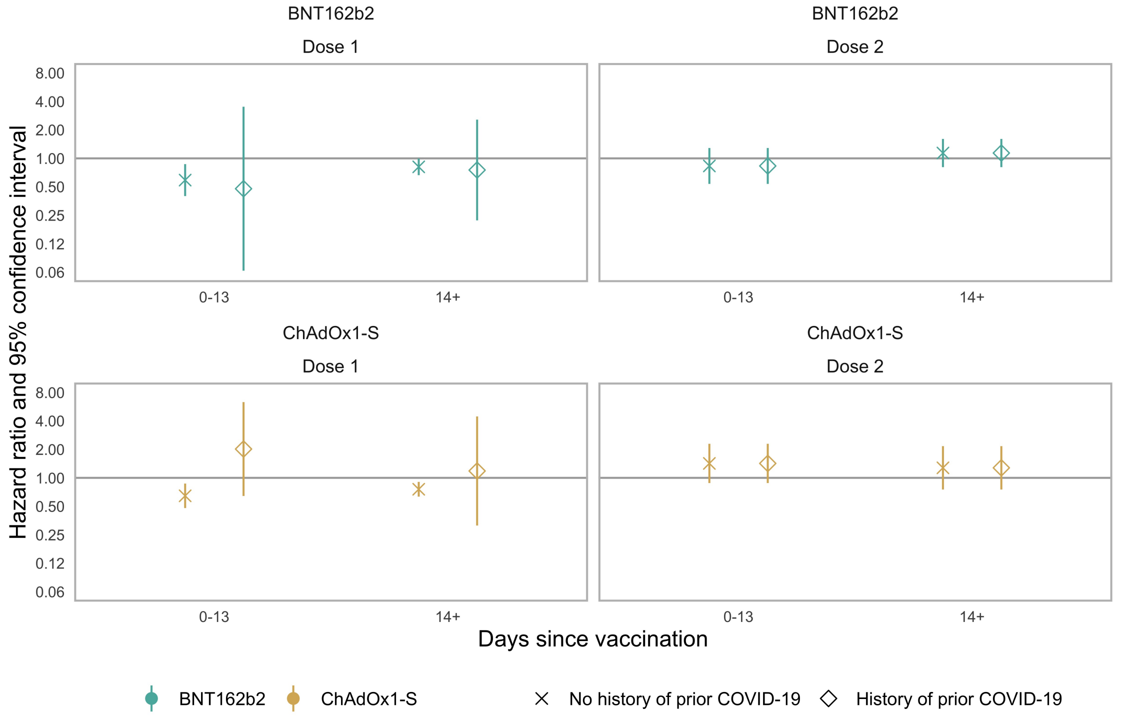


| prior_covid | dose | exposure | estimate | conf.low | conf.high | p.value |
|---|---|---|---|---|---|---|
| No | Dose 1 | BNT162b2 | 0.59 | 0.40 | 0.87 | 0.01 |
| No | Dose 1 | BNT162b2 | 0.81 | 0.66 | 1.00 | 0.05 |
| No | Dose 1 | ChAdOx1-S | 0.65 | 0.48 | 0.87 | 0.00 |
| No | Dose 1 | ChAdOx1-S | 0.76 | 0.63 | 0.91 | 0.00 |
| No | Dose 2 | BNT162b2 | 0.83 | 0.54 | 1.29 | 0.41 |
| No | Dose 2 | BNT162b2 | 1.14 | 0.81 | 1.61 | 0.46 |
| No | Dose 2 | ChAdOx1-S | 1.42 | 0.88 | 2.29 | 0.15 |
| No | Dose 2 | ChAdOx1-S | 1.28 | 0.75 | 2.16 | 0.36 |
| Yes | Dose 1 | BNT162b2 | 0.48 | 0.06 | 3.52 | 0.47 |
| Yes | Dose 1 | BNT162b2 | 0.75 | 0.22 | 2.57 | 0.65 |
| Yes | Dose 1 | ChAdOx1-S | 2.02 | 0.64 | 6.33 | 0.23 |
| Yes | Dose 1 | ChAdOx1-S | 1.18 | 0.31 | 4.46 | 0.80 |
| Yes | Dose 2 | BNT162b2 | 0.83 | 0.54 | 1.29 | 0.41 |
| Yes | Dose 2 | BNT162b2 | 1.14 | 0.81 | 1.61 | 0.46 |
| Yes | Dose 2 | ChAdOx1-S | 1.42 | 0.88 | 2.29 | 0.15 |
| Yes | Dose 2 | ChAdOx1-S | 1.28 | 0.75 | 2.16 | 0.36 |

#### Estimates, by individual outcome

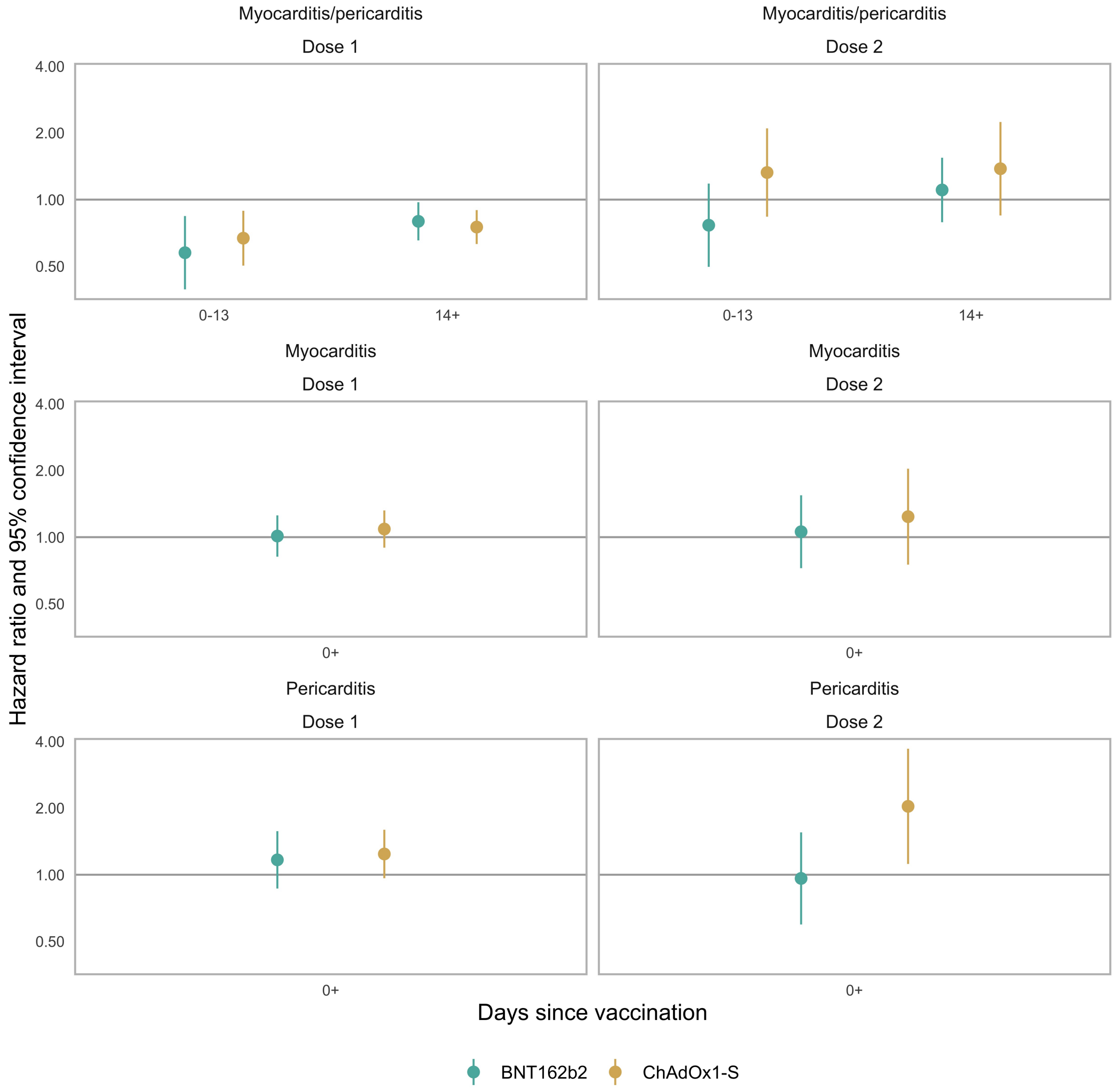


| outcome | dose | exposure | estimate | conf.low | conf.high | p.value |
|---|---|---|---|---|---|---|
| myocarditis | Dose 1 | BNT162b2 | 1.01 | 0.82 | 1.25 | 0.91 |
| myocarditis | Dose 1 | ChAdOx1-S | 1.09 | 0.90 | 1.32 | 0.39 |
| myocarditis | Dose 2 | BNT162b2 | 1.06 | 0.72 | 1.55 | 0.77 |
| myocarditis | Dose 2 | ChAdOx1-S | 1.24 | 0.75 | 2.04 | 0.40 |
| pericarditis | Dose 1 | BNT162b2 | 1.17 | 0.87 | 1.57 | 0.31 |
| pericarditis | Dose 1 | ChAdOx1-S | 1.24 | 0.96 | 1.60 | 0.09 |
| pericarditis | Dose 2 | BNT162b2 | 0.96 | 0.60 | 1.55 | 0.88 |
| pericarditis | Dose 2 | ChAdOx1-S | 2.04 | 1.12 | 3.70 | 0.02 |

#### Dose 1 replication in Wales

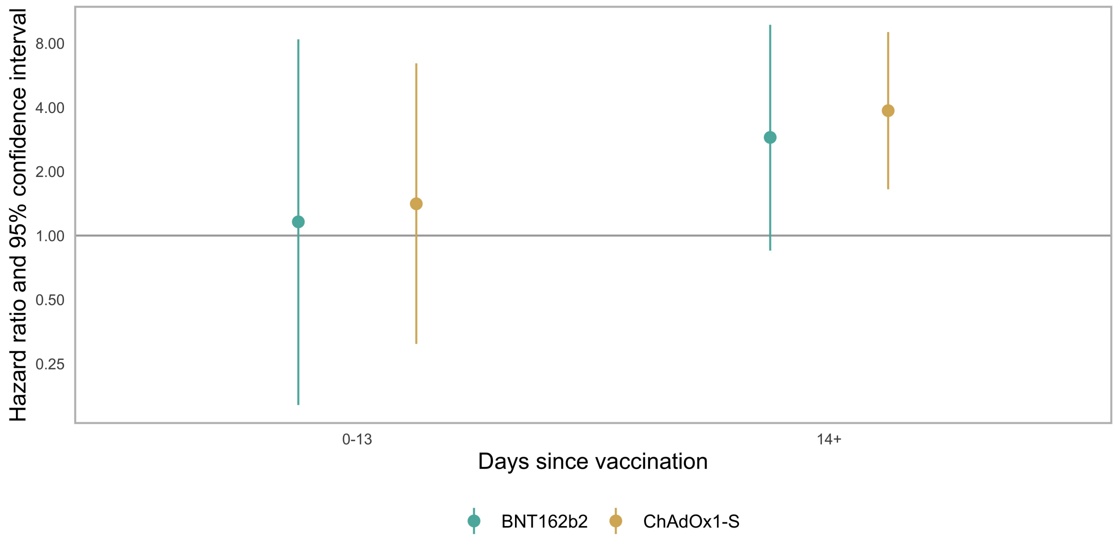


##### Events

| outcome | dose | exposure | days_post_vaccination | events |
|---|---|---|---|---|
| myocarditis/pericarditis | Dose 1 | BNT162b2 | Before | 43 |
| myocarditis/pericarditis | Dose 1 | BNT162b2 | 0-13 | <5 |
| myocarditis/pericarditis | Dose 1 | BNT162b2 | 14+ | 14 |
| myocarditis/pericarditis | Dose 1 | ChAdOx1-S | Before | 43 |
| myocarditis/pericarditis | Dose 1 | ChAdOx1-S | 0-13 | <5 |
| myocarditis/pericarditis | Dose 1 | ChAdOx1-S | 14+ | 14 |

##### Estimates

| outcome | dose | exposure | days_post_vaccination | estimate | conf.low | conf.high |
|---|---|---|---|---|---|---|
| myocarditis/pericarditis | Dose 1 | BNT162b2 | 0-13 | 1.16 | 0.16 | 8.35 |
| myocarditis/pericarditis | Dose 1 | BNT162b2 | 14+ | 2.89 | 0.85 | 9.77 |
| myocarditis/pericarditis | Dose 1 | ChAdOx1-S | 0-13 | 1.41 | 0.31 | 6.44 |
| myocarditis/pericarditis | Dose 1 | ChAdOx1-S | 14+ | 3.86 | 1.65 | 9.03 |

#### Contributions to this work

This work was conducted on behalf of the BHF Data Science Centre (Health Data Research UK) CVD-COVID-UK/COVID-IMPACT Consortium and the Longitudinal Health and Wellbeing and Data and Connectivity UK COVID-19 National Core Studies. Samantha Ip developed the analysis code and implemented the analysis in England; Fatemeh Torabi derived the dataset for Wales and implemented the analysis in Wales; and Venexia Walker derived the dataset for England, combined and visualised the results from both nations, and drafted the report. The work relied on the support of the CCU002 project group, members of which include: Angela Wood (University of Cambridge), Arun Karthikeyan Suseeladevi (University of Bristol), Ashley Akbari (Swansea University), Cathie Sudlow (BHF Data Science Centre), Emanuele Di Angelantonio (University of Cambridge), Efosa Omigie (NHS Digital), Fatemeh Torabi (Swansea University), Hoda Abbasizanjani (Swansea University), Jennifer Cooper (University of Bristol), Jonathan Sterne (University of Bristol), Rachel Denholm (University of Bristol), Rochelle Knight (University of Bristol), Sam Hollings (NHS Digital), Samantha Ip (University of Cambridge), Spencer Keene (University of Cambridge), Spiros Denaxas (University College London), Teri-Louise North (University of Bristol), Thomas Bolton (BHF Data Science Centre), Venexia Walker (University of Bristol) and William Whiteley (University of Edinburgh). The project was conceived by this group, all members of which contributed to the development of the protocol (https://github.com/BHFDSC/CCU002_03/blob/main/protocol/COVID%20vaccination%20and%20myocarditis%20and%20pericarditis.pdf). Members of the group also reviewed and discussed analysis results and their interpretation, including in the context of other relevant published studies.

#### Funding

This work was supported by: the BHF Data Science Centre led by Health Data Research UK (BHF Grant no. SP/19/3/34678); the COVID-19 Longitudinal Health and Wellbeing National Core Study funded by the Medical Research Council [MC_PC_20030; MC_PC_20059]; the Con-COV team funded by the Medical Research Council (grant number: MR/V028367/1); Health Data Research UK, which receives its core funding from the UK Medical Research Council, Engineering and Physical Sciences Research Council, Economic and Social Research Council, Department of Health and Social Care (England), Chief Scientist Office of the Scottish Government Health and Social Care Directorates, Health and Social Care Research and Development Division (Welsh Government), Public Health Agency (Northern Ireland), British Heart Foundation (BHF) and the Wellcome Trust; the Wales COVID-19 Evidence Centre, funded by Health and Care Research Wales; the ADR Wales programme of work, which is aligned to the priority themes as identified in the Welsh Government's national strategy, Prosperity for All, and brings together data science experts at Swansea University Medical School, staff from the Wales Institute of Social and Economic Research, Data and Methods (WISERD) at Cardiff University and specialist teams within the Welsh Government to develop new evidence which supports Prosperity for All by using the SAIL Databank at Swansea University, to link and analyse anonymised data. ADR Wales is part of the Economic and Social Research Council (part of UK Research and Innovation) funded ADR UK (grant ES/S007393/1). This research is part of the Data and Connectivity National Core Study, led by Health Data Research UK in partnership with the Office for National Statistics and funded by UK Research and Innovation (grant ref: MC_PC_20058). SI was funded by the International Alliance for Cancer Early Detection, a partnership between Cancer Research UK C18081/A31373, Canary Center at Stanford University, the University of Cambridge, OHSU Knight Cancer Institute, University College London and the University of Manchester. VW was supported by the Medical Research Council Integrative Epidemiology Unit at the University of Bristol [MC_UU_00011/4] and the National Core Studies, an initiative funded by UKRI, NIHR and the Health and Safety Executive.

#### Other acknowledgements

This work makes use of de-identified data held in NHS Digital's TRE for England and the SAIL Databank for Wales, made available via the BHF Data Science Centre's CVD-COVID-UK/COVID-IMPACT consortium. This work uses data provided by patients and collected by the NHS as part of their care and support. We would like to acknowledge all data providers who make health relevant data available for research.

This study makes use of anonymised data held in the Secure Anonymised Information Linkage (SAIL) Databank. This work uses data provided by patients and collected by the NHS as part of their care and support. We would also like to acknowledge all data providers who make anonymised data available for research. We wish to acknowledge the collaborative partnership that enabled acquisition and access to the de-identified data, which led to this output. The collaboration was led by the Swansea University Health Data Research UK team under the direction of the Welsh Government Technical Advisory Cell (TAC) and includes the following groups and organisations: the SAIL Databank, Administrative Data Research (ADR) Wales, Digital Health and Care Wales (DHCW), Public Health Wales, NHS Shared Services Partnership (NWSSP) and the Welsh Ambulance Service Trust (WAST). All research conducted has been completed under the permission and approval of the SAIL independent Information Governance Review Panel (IGRP) project number 0911.

#### Data availability

The data used in this study are available in NHS Digital's TRE for England, but as restrictions apply they are not publicly available (https://digital.nhs.uk/coronavirus/coronavirus-data-services-updates/trusted-research-environment-service-for-england). The CVD-COVID-UK/COVID-IMPACT programme led by the BHF Data Science Centre (https://www.hdruk.ac.uk/helping-with-health-data/bhf-data-science-centre/) received approval to access data in NHS Digital's TRE for England from the Independent Group Advising on the Release of Data (IGARD) (https://digital.nhs.uk/about-nhs-digital/corporate-information-and-documents/independent-group-advising-on-the-release-of-data) via an application made in the Data Access Request Service (DARS) Online system (ref. DARS-NIC-381078-Y9C5K) (https://digital.nhs.uk/services/data-access-request-service-dars/dars-products-and-services). The CVD-COVID-UK/COVID-IMPACT Approvals & Oversight Board (https://www.hdruk.ac.uk/projects/cvd-covid-uk-project/) subsequently granted approval to this project to access the data within the TRE for England and the Secure Anonymised Information Linkage (SAIL) Databank. The de-identified data used in this study was made available to accredited researchers only.

The data used in this study are available in the SAIL Databank at Swansea University, Swansea, UK, but as restrictions apply they are not publicly available. All proposals to use SAIL data are subject to review by an independent Information Governance Review Panel (IGRP). Before any data can be accessed, approval must be given by the IGRP. The IGRP gives careful consideration to each project to ensure proper and appropriate use of SAIL data. When access has been granted, it is gained through a privacy protecting safe haven and remote access system referred to as the SAIL Gateway. SAIL has established an application process to be followed by anyone who would like to access data via SAIL at https://www.saildatabank.com/application-process

#### Ethical approval

The North East-Newcastle and North Tyneside 2 research ethics committee provided ethical approval for the CVD-COVID-UK/COVID-IMPACT research programme (REC No 20/NE/0161).
